# Associations between cognitive function and lifestyle factors in healthy Japanese middle-aged and older adults: A cross-sectional study

**DOI:** 10.1101/2025.11.27.25340042

**Authors:** Koki Tsuda, Kotatsu Bito, Masanobu Hibi, Takahiro Ono, Hiroshi Maruyama

## Abstract

The objective of this study is to comprehensively investigate associations between cognitive function and lifestyle factors. We analyzed data from a cross-sectional study of Japanese adults that included over 1,800 variables (e.g., physical characteristics, body composition, and lifestyle habits) and cognitive function assessed using CNS Vital Signs. For participants aged 40 years or older (*n* = 710), we performed partial correlation analysis and analysis of covariance adjusted for age, sex, and years of education. Given the large number of variables, we controlled the false discovery rate within predefined data type families (real, positive, ordered categorical, categorical) using the Benjamini–Hochberg procedure; *p*-values were adjusted to *q*-values, and, for exploratory purposes, variables with *q*-values *<* 0.1 were identified. In total, 28 variables met this criterion, with particularly prominent associations for gait characteristics, vascular function, grip strength, and oral conditions, whereas blood components and other general biomarkers did not meet the threshold. These findings suggest that monitoring of identified lifestyle and physiological factors may help facilitate early detection of cognitive decline.

## Introduction

Population aging is advancing in many countries, and cognitive decline among older adults has become a major public health concern [1]. By 2050, the number of older adults living with dementia is globally projected to reach approximately 150 million, underscoring the urgent need for multifaceted strategies to address this challenge [2]. Cognitive decline is known to trigger a variety of health problems, including physical frailty, and to significantly impair quality of life [3]. In addition, dementia imposes substantial medical costs and informal caregiving burdens, potentially leading to severe economic impacts on society [4].

To prevent the onset of dementia, increasing attention has been directed toward early at-risk or prodromal stages such as mild cognitive impairment (MCI) and subjective cognitive decline (SCD) [5], [6]. MCI is characterized by mild cognitive decline that does not interfere with daily life, whereas SCD refers to a state in which objective cognitive impairment is not detectable, yet individuals subjectively perceive a decline. People with MCI or SCD are known to be at an elevated risk of developing dementia [7], [8]; however, it is also well established that appropriate interventions, such as lifestyle modifications and structured programs, can delay the progression of cognitive decline [9], [10].

Previous studies have reported that regular physical exercise [11], [12] and dietary interventions, such as adherence to a Mediterranean diet [13], [14], may slow cognitive decline. Psychological well-being also plays an important role: stress management strategies, including meditation and mindfulness, have been suggested to reduce stress and enhance cognitive performance [15]. Furthermore, active participation in social activities has been shown to provide cognitive stimulation that may help maintain brain function [16], [17]. Taken together, these findings underscore the importance of encouraging the early detection of cognitive decline and motivating individuals to engage in preventive lifestyle interventions.

A range of methods has been developed to assess cognitive decline, including cognitive tests such as the Mini-Mental State Examination [18] and Montreal Cognitive Assessment [19], detection of biomarkers such as amyloid-*β* and tau [20], and neuroimaging using positron emission tomography [21]. However, although these methods are appropriate for patients with apparent symptoms, they are not well suited for continuous, long-term monitoring of asymptomatic individuals in terms of feasibility and cost [22]. Moreover, biomarker assessment typically requires cerebrospinal fluid sampling via lumbar puncture, which is invasive [22], and brain imaging requires expensive equipment and infrastructure [23]. For these reasons, there is a growing need for more accessible, cost-effective approaches that leverage data on lifestyle factors to detect early and subtle cognitive changes at the population level [24].

In our previous work, we collected comprehensive cross-sectional data on lifestyle factors from Japanese adults [25]. This dataset includes more than 1,800 variables encompassing physical characteristics, body composition measures, and lifestyle habits, all measured in the same participants. Importantly, it also includes results from cognitive assessments performed using CNS Vital Signs (CNSVS), a convenient tool for evaluating multiple cognitive domains, including memory, attention, and executive function [26]. Analysis of these data is expected to clarify associations between cognitive function and a broader range of lifestyle and physiological variables compared with previous studies. Building on prior findings, the present study aims to investigate associations between cognitive function and a wide spectrum of lifestyle and physical parameters including physical activity, diet, sleep, stress, and social engagement in a larger and more diverse population. Finally, we aim to provide evidence-based recommendations on lifestyle factors that individuals can focus on to maintain cognitive health and promote healthy aging.

## Materials & methods

### Study Design and Participants

This study performed as exploratory analysis using data from a previously reported single-center cross-sectional observational study [25]. The protocol of the study is briefly summarized below.

The study was approved by the Institutional Review Boards (IRBs) of Kao Corporation (Tokyo, Japan; approval number K0023-2108) in October 2021. Thereafter, for data analysis, an ethics review application was submitted to the IRBs of Preferred Networks Inc. (Tokyo, Japan; approval number ET22110047) and approved in December 2022. Participants were consecutively recruited from 19 October 2021 to 4 February 2022. Recruitment was conducted through a website operated by TES Holdings (Tokyo, Japan). The inclusion criteria were: (1) Japanese men and women aged 20 years or older, (2) ability to complete questionnaires and surveys, and (3) provision of written informed consent after understanding the study procedures. The exclusion criteria were: (1) hospitalization for severe diseases (e.g., diabetes, hypertension, arteriosclerosis, heart disease, malignant tumor, Alzheimer’s disease), (2) inability to attend the clinic independently, (3) diagnosis or suspected diagnosis of dementia, (4) symptoms suggestive of COVID-19 infection, (5) use of a cardiac pacemaker, (6) current pregnancy or possible pregnancy, and (7) any other condition deemed inappropriate for participation by the principal investigator.

All participants provided written informed consent, which detailed the data elements to be used and included consent for use of anonymized data. Participants visited Ueno Asagao Clinic (Tokyo, Japan) twice at one-week intervals. All measurements were performed by trained research coordinators and physicians following standardized operating procedures. The study was conducted in accordance with the Strengthening the Reporting of Observational Studies in Epidemiology (STROBE) guidelines. The study protocol was prospectively registered at the University Hospital Medical Information Network (UMIN000045746) on October 14, 2021.

### Cognitive Function

Cognitive function was assessed using the standardized Neurocognition Index (NCI) score obtained from the CNSVS battery [26]. CNSVS is a computerized neurocognitive screening tool that evaluates multiple cognitive domains and is widely used for the assessment of cognitive function. The standardized scores are normalized for each age group based on the CNSVS normative database, with a mean of 100. Among these scores, the NCI score represents a global measure of neurocognitive function, calculated as the mean of composite memory, psychomotor speed, reaction time, complex attention, and cognitive flexibility.

### Physical Characteristics, Body Composition, and Lifestyle Habits

Height was measured using a standard stadiometer, weight was measured using a body composition analyzer (InBody 770K, InBody Corp., Seoul, Korea), and body mass index (BMI) was calculated. Blood pressure (systolic and diastolic), cardio-ankle vascular index (CAVI), ankle-brachial index (ABI), and other vascular parameters were measured using a VS-2500 Vascular Screening System (Fukuda Denshi Co., Ltd., Tokyo, Japan). Grip strength was measured twice with a Smedley-type dynamometer (GRIP-D, Takei Scientific Instruments Co., Ltd., Tokyo, Japan), and the mean value was recorded.

Gait parameters including walking speed, stride length, step width, foot angle, stance and swing phases, and plantar pressure distribution were measured on a 6.4-m walkway equipped with a sheet-type foot pressure sensor (Anima Corp., Tokyo, Japan); the mean of four trials was used for analysis. Daily walking speed and step counts were recorded continuously for 14 days using a tri-axial accelerometer (HW-100, Kao Corporation, Tokyo, Japan) and a smartphone-based gait analysis application (Chami, InfoDeliver Co., Ltd., Tokyo, Japan).

Information on medical history, medication use, and lifestyle factors, including diet, sleep, psychological health, stress, excretory function, and menopausal status, was obtained through physician or coordinator interviews and a series of validated questionnaires: the brief-type self administered diet history questionnaire (BDHQ) [27], International Physical Activity Questionnaire (IPAQ) [28], [29], Questionnaire for Medical Checkup of Old-Old (QMCOO) [30], dietary habits questionnaire [31], Athens Insomnia Scale [32], [33], Oguri–Shirakawa–Azumi Sleep Inventory MA version (OSA-MA) [34], Berlin Questionnaire [35], World Health Organization–Five Well-Being Index (WHO-5) [36], Brief Job Stress Questionnaire (BJSQ) [37], Chalder Fatigue Scale [38], Center for Epidemiologic Studies Depression Scale (CES-D) [39], [40], Fatigue Feelings Questionnaire [41], Sun Exposure Questionnaire [42], Ten-Item Personality Inventory (TIPI-J) [43], [44], Oxford Happiness Questionnaire [45], Overactive Bladder Symptom Score (OABSS) [46], International Consultation on Incontinence Questionnaire–Urinary Incontinence Short Form (ICIQ-UI SF) [47], Fecal Incontinence Quality of Life (FIQL) scale [48], Neurogenic Bowel Dysfunction (NBD) score [49], Kupperman Menopausal Index [50], modified Menstrual Distress Questionnaire (mMDQ) [51], [52], climacteric and senescence scores [53], and, for participants within 3 years postpartum, the Edinburgh Postnatal Depression Scale (EPDS) [54]. Oral health indicators, including tooth and gum condition, halitosis, and oral dryness were assessed using a five-point rating questionnaire developed specifically for this study.

In addition, various physiological and biochemical parameters were measured using the methods described in protocol [25] and included in the analyses. Assessments included physical performance tests, laboratory analysis of blood, urine, and saliva, an oral glucose tolerance test, liquid chromatography–tandem mass spectrometry analysis of chiral amino acids, lipid mediators, vitamin D metabolites, and polyamines, assessment of hair loss, analysis of hand surface characteristics, analysis of stratum corneum lipids and sebum, analysis of body odor, skin surface spectroscopy, and analysis of RNA from skin surface lipids (SSL-RNA).

### Statistical Analyses

The data snapshot used for the analyses were downloaded on 23 July 2024. Only records that did not contain any personally identifiable information were used for analyses, and investigators had no access to such information.

The dataset included over 1,800 variables describing anthropometric measures, body composition, and lifestyle factors. To identify lifestyle factors associated with cognitive function, we performed exploratory statistical analyses. Because sex, age, and years of education are widely recognized as major determinants of cognitive performance [55], [56], [57], we calculated partial correlation coefficients and performed analysis of covariance (ANCOVA) to control for their potential confounding effects. To address the issue of multiple comparisons, *p*-values were adjusted using the Benjamini–Hochberg procedure [58] to yield *q*-values, thereby controlling the false discovery rate (FDR). For exploratory purposes, statistical significance was defined as *q*-values < 0.1. All statistical analyses were conducted in Python (v3.9.20) using the pingouin (v0.5.5), statsmodels (v0.14.4), and SciPy (v1.13.1) packages. Each variable was assumed to be one of four data types: real (continuous real-valued), positive (continuous and strictly positive), ordered categorical (ordinal), or categorical (nominal) [59]. Methods for handling missing values and statistical tests were applied accordingly.

### Treatment of Missing Data

The dataset contained missing values in approximately 0.2%–57% of variables [59]. Appropriate handling of missing data is essential for valid statistical inference [60]; however, given the large number of lifestyle-related variables, it was not feasible to apply tailored imputation methods for each variable while accounting for inter-variable relationships. Therefore, we adopted a complete-case analysis approach, excluding participants with missing data on the variables included in each analysis. We acknowledge that this approach may introduce bias into the estimated parameters.

### Analysis of Real, Positive, and Ordered Categorical Variables

For variables classified as the real, positive, or ordered categorical type, we calculated partial Spearman correlation coefficients between each variable and the NCI score, adjusting for sex, age, and years of education, and obtained *p*-values. Only participants with complete data for the variables were included in this analysis. Cognitive subdomain scores other than the NCI score were excluded because they were expected to be highly correlated with the NCI score. After applying FDR correction within each data type, variables with *q*-values < 0.1 were identified for further interpretation.

### Analysis of Categorical Variables

For the categorical type variables, ANCOVA was performed with the NCI score as the dependent variable and sex, age, and years of education as covariates. Only participants with complete data for the variables were included in this analysis. Continuous variables were standardized, and categorical variables were converted into dummy variables before inclusion in the model. As a preliminary assumption check, interaction terms between each categorical variable and the covariates were included in the model to assess homogeneity of regression slopes; variables with non-significant interaction terms (*p*-values > 0.05) were considered to satisfy this assumption. Homogeneity of variance was further assessed using Levene’s test (*p*-values > 0.05), and variables that did not meet these assumptions were excluded from the subsequent ANCOVA. After applying FDR correction, variables with *q*-values < 0.1 were identified for further interpretation.

## Results

### Participants

The dataset included 997 Japanese adults. After excluding three individuals who did not provide consent for secondary use of data, 994 participants remained. Among these, 715 participants aged 40 years or older, who constituted the target population for examining potential age-related cognitive decline, were selected. After excluding five participants with missing NCI scores, a total of 710 participants were included in the final analysis. The participant selection flow is illustrated in Fig 1. Summary statistics of the demographic and clinical characteristics of the analyzed participants are presented in Table 1.

**Table 1.** Detailed demographic, lifestyle, and cognitive characteristics of participants included in the final analysis (*n* = 710).

| Variables | *n* | Percent (%) |
| --- | --- | --- |
| **Gender** |  |  |
| Male | 349 | 49.2 |
| Female | 361 | 50.8 |
| **Age ($59.3 \pm 11.2$ [40–79])** |  |  |
| $40 \leq \text{to} < 50$ | 183 | 25.8 |
| $50 \leq \text{to} < 60$ | 164 | 23.1 |
| $60 \leq \text{to} < 70$ | 161 | 22.7 |
| $70 \leq \text{to} < 80$ | 202 | 28.5 |
| **BMI ($22.8 \pm 3.7$ [13.5–41.0])** |  |  |
| Underweight ($< 18.5$) | 79 | 11.1 |
| Normal range ($18.5 \leq \text{to} < 25$) | 459 | 64.6 |
| Pre-obese ($25 \leq \text{to} < 30$) | 142 | 20.0 |
| Obese class I ($30 \leq \text{to} < 35$) | 26 | 3.7 |
| Obese class II ($35 \leq \text{to} < 40$) | 2 | 0.3 |
| Obese class III ($\geq 40$) | 2 | 0.3 |
| **Years of education ($15.0 \pm 2.3$ [6–30])** |  |  |
| $\leq 9$ | 15 | 2.1 |
| $9 < \text{to} \leq 12$ | 132 | 18.6 |
| $12 < \text{to} \leq 16$ | 480 | 67.6 |
| $16 < \text{to} \leq 18$ | 57 | 8.0 |
| $18 < \text{to} \leq 21$ | 20 | 2.8 |
| $> 21$ | 6 | 0.8 |
| **Smoking status** |  |  |
| Never smoker | 411 | 57.9 |
| Former smoker | 206 | 29.0 |
| Current smoker (1–14 cigarettes per day) | 50 | 7.0 |
| Current smoker (15–29 cigarettes per day) | 40 | 5.6 |
| Current smoker ($\geq 30$ cigarettes per day) | 3 | 0.4 |
| **Drinking frequency** |  |  |
| Every day | 148 | 20.8 |
| Sometimes | 292 | 41.1 |
| Seldom | 270 | 38.0 |
| **Exercise frequency** |  |  |
| Almost never | 277 | 39.0 |
| Once per month | 17 | 2.4 |
| Two or three times per month | 31 | 4.4 |
| Once per week | 77 | 10.8 |
| Two to four times per week | 161 | 22.7 |
| Five or six times per week | 88 | 12.4 |
| Every day | 59 | 8.3 |
| **NCI score ($102.4 \pm 10.7$ [48–124])** |  |  |
| - | 710 | 100.0 |

Continuous variables are expressed as mean ± standard deviation [range]. BMI, body mass index [ $\text{kg}/\text{m}^2$ ]; NCI, Neurocognition Index.

**Figure 1.**
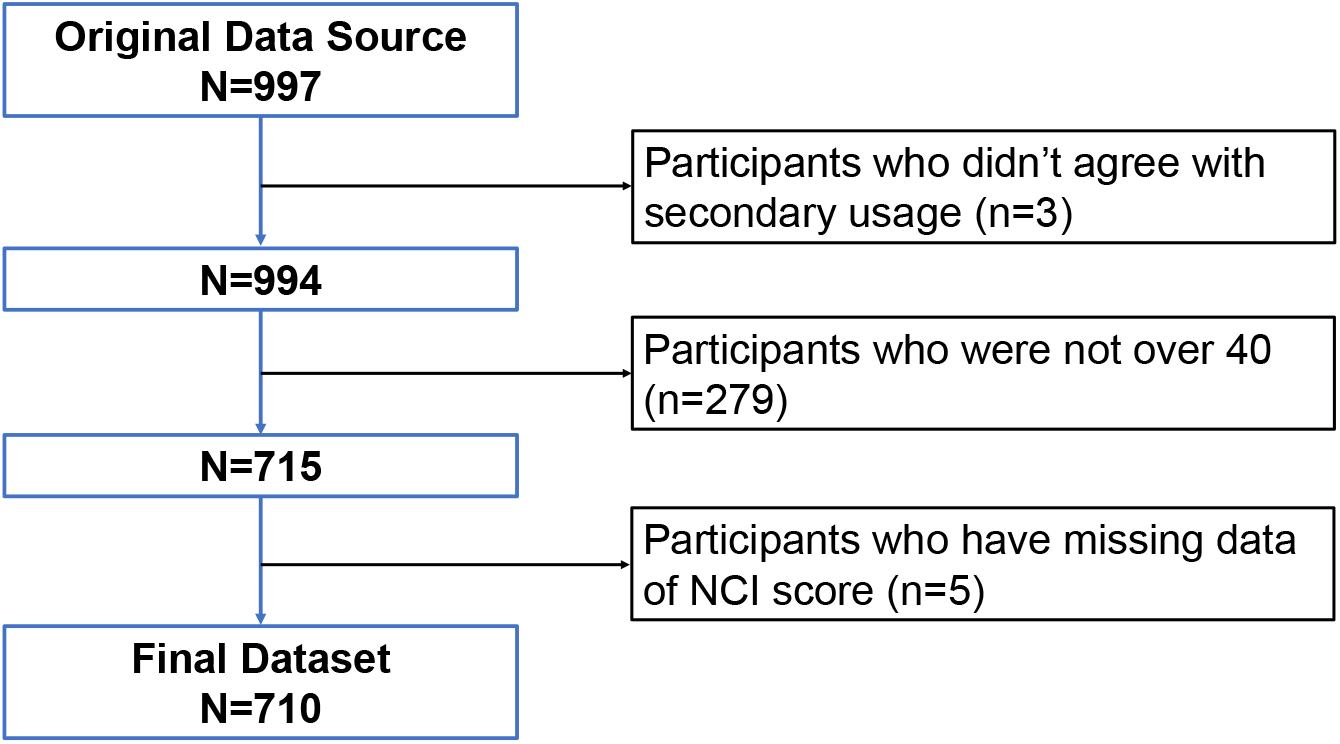
Participant selection flow. A flow diagram showing the inclusion and exclusion process of study participants. Among the 997 initially recruited Japanese adults, three individuals who did not provide consent for secondary use of data, 279 participants younger than 40 years, and five participants with missing NCI scores were excluded, resulting in 710 participants included in the final analysis.

The final dataset included 349 men (49.2%) and 361 women (50.8%), with a mean age of 59.3 ± 11.2 years, mean BMI of 22.8 ± 3.7 kg/m^2^, and mean years of education of 15.0 ± 2.3. Most participants were either never-smokers (*n* = 411, 57.9%) or former smokers (*n* = 206, 29.0%). Regarding alcohol consumption, the majority reported drinking occasionally (*n* = 292, 41.1%) or rarely (*n* = 270, 38.0%). With respect to exercise habits, most participants reported either little to no regular exercise (*n* = 277, 39.0%) or exercising two to four times per week (*n* = 161, 22.7%). The mean NCI score was 102.4 ± 10.7.

### Analyses of associations with cognitive function

The results of the association analyses are presented in Tables 2 and 3 and Figs 2 and 3. From the partial correlation analysis of the real type variables, many gait related variables measured using a sheet-type pressure sensor or a smartphone application were associated with cognitive function. For example, increases in double support phase, stance phase, and gait cycle, as well as decreases in walking speed, cadence (step frequency, expressed as steps per minute), and swing phase, were associated with lower cognitive function. In addition, a higher knee pain score and a lower activities of daily living (ADLs) score, derived from gait characteristics, were both associated with lower cognitive function. Overall, participants with slower walking speed and longer gait cycle tended to exhibit lower cognitive function.

**Table 2.** Associations between NCI scores and the real type variables.

| Variables | Category Field | *n* | Partial *r* | 95% CI | *p*-value | *q*-value (FDR) |
| --- | --- | --- | --- | --- | --- | --- |
| L-tb | Vascular function | 708 | -0.148 | [-0.22, -0.08] | < 0.001 | 0.0614 |
| Right double support phase | Walking characteristics | 710 | -0.141 | [-0.21, -0.07] | < 0.001 | 0.0614 |
| Left double support phase | Walking characteristics | 710 | -0.140 | [-0.21, -0.07] | < 0.001 | 0.0614 |
| Mean walking speed (segments ≥ 20 m) | Walking characteristics | 566 | 0.140 | [0.06, 0.22] | < 0.001 | 0.0837 |
| Right stance phase | Walking characteristics | 710 | -0.138 | [-0.21, -0.06] | < 0.001 | 0.0614 |
| ACOT2 (SSL-RNA, RPM correction) | Biomarker | 549 | 0.137 | [0.05, 0.22] | 0.00138 | 0.0892 |
| Left stance phase | Walking characteristics | 710 | -0.136 | [-0.21, -0.06] | < 0.001 | 0.0614 |
| Mean walking speed (smartphone app.) | Walking characteristics | 551 | 0.133 | [0.05, 0.21] | 0.00187 | 0.0906 |
| Knee pain score | Walking characteristics | 709 | -0.131 | [-0.20, -0.06] | < 0.001 | 0.0830 |
| Right relative stance phase | Walking characteristics | 709 | -0.128 | [-0.20, -0.05] | < 0.001 | 0.0837 |
| Right relative swing phase | Walking characteristics | 709 | 0.126 | [0.05, 0.20] | < 0.001 | 0.0837 |
| Cadence (step frequency) (AVM method) | Walking characteristics | 701 | 0.125 | [0.05, 0.20] | < 0.001 | 0.0837 |
| KRT79 (SSL-RNA, RPM correction) | Biomarker | 614 | 0.125 | [0.05, 0.20] | 0.00201 | 0.0906 |
| ADLs | Walking characteristics | 710 | 0.124 | [0.05, 0.20] | < 0.001 | 0.0837 |
| Preferred walking speed (pressure sensor) | Walking characteristics | 710 | 0.124 | [0.05, 0.20] | < 0.001 | 0.0837 |
| Preferred walking speed (AVM method) | Walking characteristics | 701 | 0.123 | [0.05, 0.20] | 0.00109 | 0.0868 |
| L-ABI | Vascular function | 708 | 0.121 | [0.05, 0.19] | 0.00127 | 0.0892 |
| Left grip strength | Motor function | 710 | 0.120 | [0.05, 0.19] | 0.00137 | 0.0892 |
| Right gait cycle | Walking characteristics | 709 | -0.119 | [-0.19, -0.05] | 0.00159 | 0.0906 |
| Cadence (step frequency) | Walking characteristics | 709 | 0.118 | [0.04, 0.19] | 0.00165 | 0.0906 |
| Left stance phase (AVM method) | Walking characteristics | 701 | -0.118 | [-0.19, -0.04] | 0.00177 | 0.0906 |
| Gait-derived age | Walking characteristics | 710 | -0.118 | [-0.19, -0.04] | 0.00174 | 0.0906 |
| Left gait cycle | Walking characteristics | 709 | -0.117 | [-0.19, -0.04] | 0.00193 | 0.0906 |
| Right grip strength | Motor function | 709 | 0.115 | [0.04, 0.19] | 0.00220 | 0.0948 |

This table lists the real type variables that exhibited associations with cognitive function after adjustment for sex, age, and years of education. The variables are sorted by the absolute value of the partial correlation coefficient. *q*-values were adjusted for multiple testing using FDR method. NCI, Neurocognition Index; FDR, false discovery rate; CI, confidence interval; SSL-RNA, RNA from skin surface lipids; RPM, reads per million; AVM, around view monitor; ADLs, activities of daily living; L-ABI, left ankle-brachial index. (See the details in S1 Table.)

**Table 3.** Associations between NCI scores and the categorical type variables.

| Variables | Category Field | *n* | df | Partial $\eta_p^2$ | F-value | *p*-value | *q*-value (FDR) |
| --- | --- | --- | --- | --- | --- | --- | --- |
| Medical history pneumothorax | Medical history / medication | 710 | 1 | 0.0380 | 27.81 | < 0.001 | < 0.001 |
| Dry mouth | Oral cavity | 710 | 5 | 0.0334 | 4.85 | < 0.001 | 0.0390 |
| Jaw pain | Oral cavity | 710 | 5 | 0.0300 | 4.34 | < 0.001 | 0.0721 |
| Taste impairment | Oral cavity | 710 | 4 | 0.0299 | 5.40 | < 0.001 | 0.0390 |

This table lists the categorical type variables that exhibited associations with cognitive function in the ANCOVA after adjustment for sex, age, and years of education. Variables are sorted by the partial $\eta_p^2$ values. *q*-values were adjusted for multiple testing using the FDR method. NCI, Neurocognition Index; ANCOVA, analysis of covariance; FDR, false discovery rate; df, degrees of freedom. “Dry mouth”, “Jaw pain”, and “Taste impairment” were assessed using the oral hygiene questionnaire. (See the details in S2 Table.)

**Figure 2.**
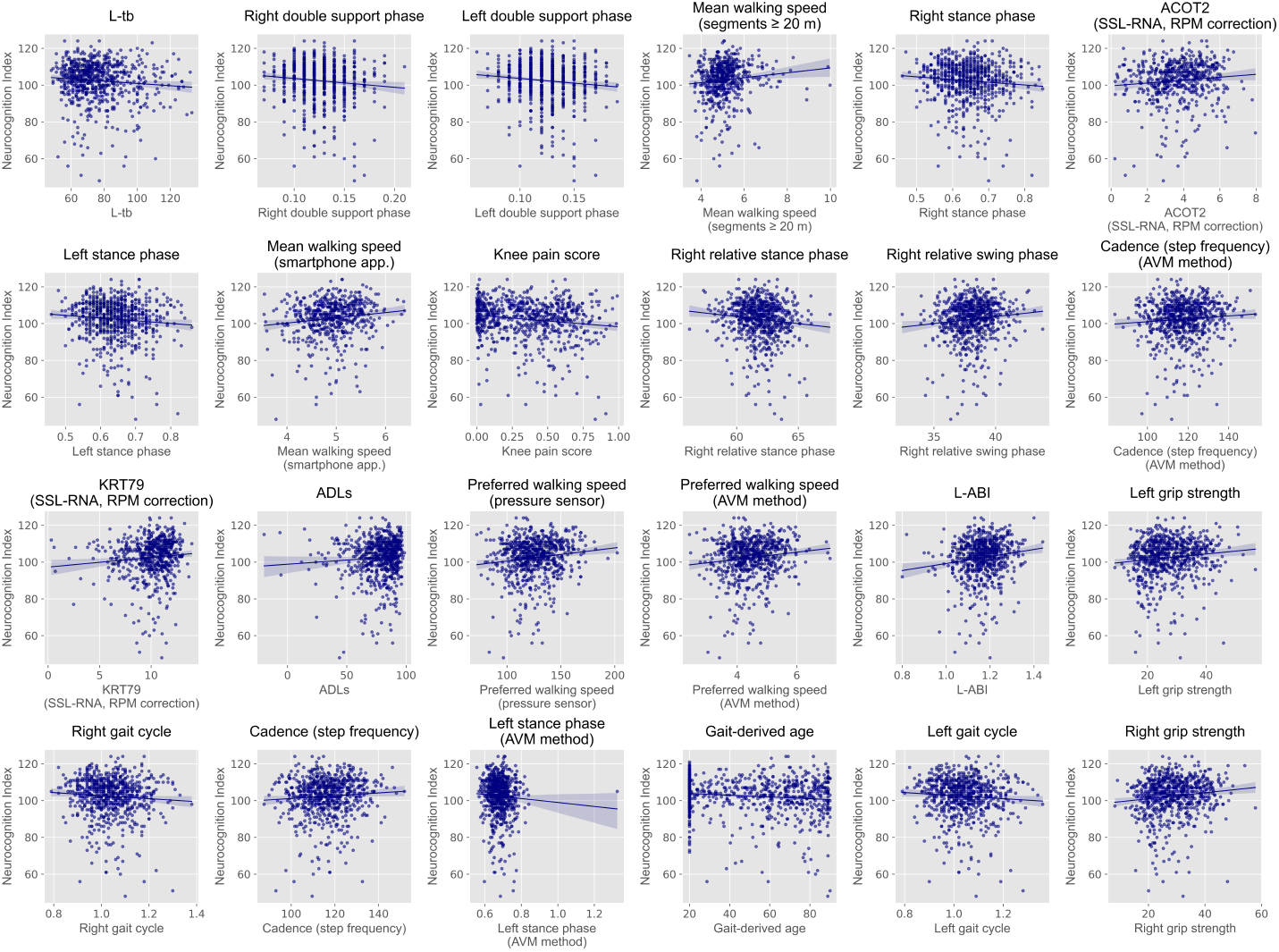
Scatter plots of NCI scores against the real type variables identified in partial correlation analysis. Each plot shows the relationship between NCI scores and the real type variables that exhibited partial correlations with cognitive function after adjustment for sex, age, and years of education (*q*-values < 0.1). Lines represent linear regression fits with 95% confidence intervals. NCI, Neurocognition Index.

**Figure 3.**
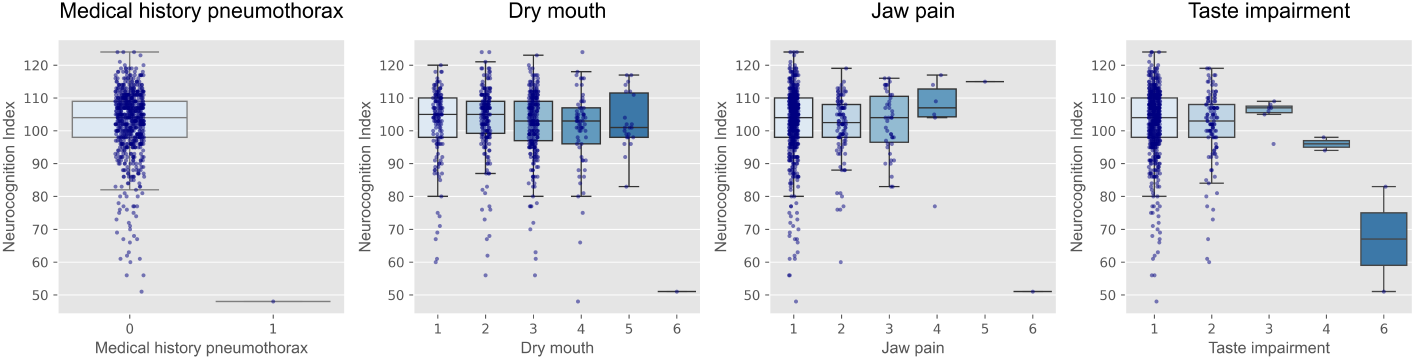
Box plots of NCI scores across the categorical type variables identified in ANCOVA. Each panel shows the distribution of NCI scores according to the categorical type variables that exhibited associations with cognitive function (*q*-values < 0.1) in the ANCOVA after adjustment for sex, age, and years of education. Variables include medical history of pneumothorax from the medical history questionnaire and oral conditions from the oral hygiene questionnaire. Boxes represent interquartile ranges, horizontal lines indicate medians, and dots represent individual participants. NCI, Neurocognition Index; ANCOVA, analysis of covariance.

Among vascular function related variables, a decrease in L-ABI indicating a higher degree of vascular stenosis in the left leg was also associated with lower cognitive function. Furthermore, among motor function related variables, lower grip strength in both hands was associated with lower cognitive function.

From the ANCOVA results for the categorical variables, several oral health related variables were identified. Specifically, increased oral dryness and decreased sense of taste were associated with cognitive decline. Additionally, relationships were observed between cognitive function and several other factors, including L-tb (a pulse wave parameter), RNA expression levels in skin-surface lipids, and a medical history of pneumothorax. No variables with positive and ordered categorical type showed significant associations after FDR correction (*q*-values < 0.1).

## Discussion

In this study, we conducted a comprehensive statistical analysis of a dataset comprising anthropometric characteristics, body composition, and lifestyle habits of Japanese adults aged 40 years or older, with the aim of exploring factors associated with cognitive function. After adjusting for sex, age, and years of education, we conducted partial correlation analysis and ANCOVA, which revealed that variables related primarily to gait characteristics, vascular function, motor function, and oral conditions were associated with cognitive function. By contrast, no significant associations were found for common biochemical markers in this analysis. These findings suggest that lifestyle related indicators may serve as useful markers for monitoring cognitive decline.

Our finding that quantitative gait characteristics are associated with cognitive function is consistent with previous studies [61], [62]. Gait is a complex motor activity that relies on multiple cognitive domains, including attention, planning, visuospatial processing, and motor control [63]. Therefore, gait may serve as sensitive indicators of cognitive decline. These findings suggest that daily gait monitoring, using smartphones or wearable devices, could provide a useful approach for early detection of cognitive decline in adults aged 40 years or older.

ABI is a well-established indicator for assessing the degree of vascular stenosis or occlusion in the lower limbs due to atherosclerosis, and it is widely employed as a non-invasive screening tool for peripheral artery disease [64]. Our results, showing lower cognitive function among individuals with reduced ABI values (indicating greater vascular stenosis), are in agreement with previous reports [65]. Vascular narrowing leads to chronic cerebral hypoperfusion, which has been associated with reduced oxygen supply, neurodegeneration, and impaired clearance of amyloid-*β* [66], [67]. These changes are thought to contribute to cognitive decline.

Handgrip strength is a well-recognized biomarker of physical function and aging, and its association with cognitive function has been reported in many studies [68], [69], [70]. Several mechanisms have been proposed to explain this relationship. For instance, muscle weakness and cognitive impairment share common factors such as high levels of inflammatory markers and reduced sex steroid levels [71], [72]. Muscle strength also serves as a general indicator of central nervous system integrity, and decreased muscle strength may reflect an overall reduction in neural processing activity, consequently leading to impairments across various cognitive domains [73], [74]. Although the present dataset included several body composition variables that reflect total muscle mass, no significant association with cognitive function was observed. This may be because muscle mass does not necessarily reflect muscle strength, whereas grip strength which requires fine and coordinated movements of the hand and forearm may better capture neuromuscular efficiency. This could explain the stronger association between grip strength and cognitive function.

The relationship between oral conditions and cognitive function has been increasingly reported in recent years [75], [76], [77]. Tooth loss, for example, leads to reduced masticatory function and has been associated with a decrease in hippocampal neuron numbers, reduced central nervous system activity, and reduced brain volume [78]. Salivary hyposecretion has been implicated in xerostomia (dry mouth), taste disorders, and periodontal diseases [79], [80]. In a rat model study, periodontitis was reported to elevate inflammatory cytokine levels in the brain, which may be implicated in the pathogenesis of dementia [81]. Variables related to the oral conditions identified in our study may reflect these pathophysiological processes.

Additional associations were found between cognitive function and the pulse wave parameter, consistent with prior studies reporting that faster pulse wave velocity is associated with lower cognitive function [82], [83]. Because pulse wave velocity is an established indicator of arteriosclerosis [84], this finding supports the proposed association between ABI and cognitive decline. We also observed associations with certain RNA biomarkers derived from skin surface and a history of pneumothorax, although these associations have not been previously reported, and their underlying mechanisms remain unclear.

Taken together, these findings from our comprehensive statistical analysis demonstrated that variables related to gait, vascular function, grip strength, and oral condition were associated with cognitive function in Japanese adults aged 40 years or older. Nearly all observed associations were generally supported by prior studies. Importantly, even after adjusting for age, sex, and years of education, lifestyle-related variables such as gait characteristics and oral conditions were identified more frequently than biochemical or hematological variables, underscoring their potential relevance as early indicators of cognitive decline. Furthermore, these indicators can be non-invasively monitored using accelerometers or questionnaires, suggesting their potential utility for early detection of cognitive decline and the prevention of dementia.

### Limitations

This study has several limitations. First, there is a potential for bias arising from the exclusion of participants with missing data. As mentioned earlier, we calculated the statistics after listwise deletion of missing values. If the missingness occurred according to a specific pattern, the resulting statistical analyses may be biased. Second, this study does not conclude that variables that did not show an association with cognitive function are truly unrelated to cognitive function. The absence of a detected association does not imply the absence of a true relationship. Considering these limitations, further studies are warranted to elucidate the mechanisms and potential causal relationships underlying the findings of this study.

## Conclusion

In this study, we analyzed data on 710 Japanese adults aged 40 years or older from a multiparameter cross-sectional study conducted in 2021-2022. Using partial correlation analysis and analysis of covariance adjusted for age, sex, and years of education, we examined associations between cognitive function and more than 1,800 variables encompassing physical characteristics, body composition, and lifestyle habits. We identified 28 variables related to gait, vascular function, grip strength, and oral condition that were associated with cognitive function as assessed using CNS Vital Signs. These findings suggest that appropriate monitoring of such lifestyle-related indicators may enable early detection of cognitive decline and contribute to preventive strategies for various cognitive disorders.

## Data Availability

Owing to ethical and contractual restrictions, the underlying individual-level data cannot be made publicly available. Researchers who wish to request access to the data should contact the Digital Business Creation division at Kao Corporation. Requests will be considered in consultation with the relevant ethical committees and IRBs, and access may be granted subject to appropriate agreements and approvals.

## Acknowledgments

The authors thank Yuki Saito, Aya Kawakami, Shun Katada, and Aiko Suzuki for their valuable contributions to the design and execution of the cross-sectional study.

We also gratefully acknowledge Kei Sugitani, Adeline Muliandi, Nami Yamanaka, Takahiro Hasumura, Yasutoshi Ando, Takashi Fushimi, Teruhisa Fujimatsu, Tomoki Akatsu, Sawako Kawano, Ren Kimura, Shigeki Tsuchiya, Yuuki Yamamoto, Mai Haneoka, Ken Kushida, Tomoki Hideshima, Eri Shimizu, Jumpei Suzuki, Aya Kirino, Hisashi Tsujimura, Shun Nakamura, Takashi Sakamoto, Yuki Tazoe, Masayuki Yabuki, Shinobu Nagase, Tamaki Hirano, Reiko Fukuda, Yukari Yamashiro, Yoshinao Nagashima, Nobutoshi Ojima, Motoki Sudo, Naoki Oya, Yoshihiko Minegishi, and Koichi Misawa for conducting the instrumental and biochemical analyses to identify the numerous health variables across diverse categories.

## Supplementary

S1 Table.

### Detailed descriptions of the real type variables associated with cognitive function

This supplementary table provides detailed definitions and measurement descriptions of the real type variables that exhibited partial correlations with Neurocognition Index (NCI) scores after adjustment for sex, age, and years of education.

S2 Table.

### Definition of the categorical type variables associated with cognitive function

This supplementary table provides detailed definitions and response options for the categorical type variables that showed associations with Neurocognition Index (NCI) scores in the analysis of covariance (AN-COVA) adjusted for age, sex, and years of education.

