## Supplementary material for "Associations between cognitive function and lifestyle factors in healthy Japanese middle-aged and older adults: A cross-sectional study": S1 Table

**S1 Table. Detailed descriptions of the real type variables associated with cognitive function.** This supplementary table provides detailed definitions and measurement descriptions of the real type variables that exhibited partial correlations with Neurocognition Index (NCI) scores after adjustment for sex, age, and years of education.
Each variable was assigned to one of the predefined field categories (e.g., Walking characteristics,　Vascular function) based on the nature of the measurement. Variables were derived from standardized physical, biochemical, and digital assessments conducted in the cross-sectional study.

| **Variable name** | **Category field** | **Description / definition** | **Unit (source)** |
| --- | --- | --- | --- |
| **L-tb** | Vascular function | Time from the first component of the second heart sound to the dicrotic notch of the left brachial pulse wave. | ms (PWV device) |
| **Right double support phase** | Walking characteristics | Percentage of gait cycle during which both feet are on the ground (right side) during a standard walking test. | % (pressure sensor) |
| **Left double support phase** | Walking characteristics | Same as above for the left side. | % (pressure sensor) |
| **Mean walking speed (segments ≥ 20 m)** | Walking characteristics | Mean speed across multiple habitual walking sessions (≥ 20 m) recorded by smartphone. | m/s (app) |
| **Right stance phase** | Walking characteristics | Proportion of gait cycle spent with the right foot in contact with the ground during a standard walking test. | % (pressure sensor) |
| **ACOT2 (SSL-RNA, RPM correction)** | Biomarker | mRNA expression level of acyl-CoA thioesterase 2 in skin surface lipids (RNA-seq). | normalized RPM |
| **Left stance phase** | Walking characteristics | As above for the left side during a standard walking test. | % (pressure sensor) |
| **Mean walking speed (smartphone app.)** | Walking characteristics | Mean speed across multiple habitual walking sessions recorded by smartphone. | m/s |
| **Knee pain score** | Walking characteristics | Self-reported pain level estimated from gait-derived features using machine-learning model. | arbitrary units |
| **Right relative stance phase** | Walking characteristics | Ratio of stance duration to total gait cycle for right side during a standard walking test. | % (pressure sensor) |
| **Right relative swing phase** | Walking characteristics | Complementary ratio to stance phase for right side during a standard walking test. | % (pressure sensor) |
| **Cadence (step frequency) (AVM method)** | Walking characteristics | Steps per minute calculated using the around-view monitoring (AVM) method during a standard walking test. | steps/min |
| **KRT79 (SSL-RNA, RPM correction)** | Biomarker | mRNA expression level of keratin 79 in skin surface lipids (RNA-seq). | normalized RPM |
| **ADLs** | Walking characteristics | Activities of daily living score estimated from multi-dimensional gait parameters during a standard walking test. | score (0–100) |
| **Preferred walking speed (pressure sensor)** | Walking characteristics | Preferred (self-selected) walking speed measured during a standard walking test on pressure mat. | m/s |
| **Preferred walking speed (AVM method)** | Walking characteristics | Preferred (self-selected) walking speed derived from 3D body-tracking system (around-view monitoring) during a standard walking test. | m/s |
| **L-ABI** | Vascular function | Left ankle–brachial index (ankle systolic pressure / arm systolic pressure). | ratio |
| **Left grip strength** | Motor function | Maximal voluntary grip force of left hand measured by dynamometer. | kg |
| **Right gait cycle** | Walking characteristics | Duration of a full gait cycle (right side) during a standard walking test. | s |
| **Cadence (step frequency)** | Walking characteristics | Habitual steps per minute based on accelerometry. | steps/min |
| **Left stance phase (AVM method)** | Walking characteristics | Stance phase duration from around-view motion capture during a standard walking test. | % |
| **Gait-derived age** | Walking characteristics | Estimated biological age from multi-dimensional gait parameters during a standard walking test. | years (model-derived) |
| **Left gait cycle** | Walking characteristics | Duration of a full gait cycle (left side) during a standard walking test. | s |
| **Right grip strength** | Motor function | Maximal voluntary grip force of right hand measured by dynamometer. | kg |
