## Supplementary material for "Associations between cognitive function and lifestyle factors in healthy Japanese middle-aged and older adults: A cross-sectional study": S2 Table

**S2 Table. Detailed descriptions of the categorical type variables associated with cognitive function.** This supplementary table provides detailed definitions and response options for the categorical type variables that showed associations with Neurocognition Index (NCI) scores in the analysis of covariance (ANCOVA) adjusted for age, sex, and years of education.

Each variable was assigned to one of the predefined field categories (e.g., Oral cavity) based on the nature of the variables. Variables were derived from the medical history questionnaire and the oral hygiene questionnaire administered in the cross-sectional study.

| **Variable name** | **Category field** | **Description** | **Response options (value labels)** |
| --- | --- | --- | --- |
| **Medical history pneumothorax** | Medical history / medication | Self-reported history of pneumothorax. Participants were asked whether they had ever been diagnosed with pneumothorax. | 0 = No (no history); 1 = Yes (history present) |
| **Dry mouth** | Oral cavity | Frequency of perceived oral dryness during the past 2–3 months was assessed using the following question:  “Do you feel that your mouth is dry?” | 1 = Never; 2 = Rarely; 3 = Sometimes; 4 = Often; 5 = Always; 6 = Do not know |
| **Jaw pain** | Oral cavity | Frequency of pain in the jaw area during the past 2–3 months was assessed using the following question:  “Do you experience pain in your jaw?” | 1 = Never; 2 = Rarely; 3 = Sometimes; 4 = Often; 5 = Always; 6 = Do not know |
| **Taste impairment** | Oral cavity | Degree of difficulty perceiving taste during the past 2–3 months was assessed using the following question:  “Do you have difficulty perceiving taste?” | 1 = Never; 2 = Rarely; 3 = Sometimes; 4 = Often or Always; 6 = Do not know |
